# Impact of meteorological parameters on the Covid-19 incidence. The case of the city of Oran, Algeria

**DOI:** 10.1101/2020.07.10.20151258

**Authors:** Farid Rahal, Salima Rezak, Fatima Zohra Baba Hamed

**Affiliations:** Architecture department, Laboratoire des Sciences, Technologie et Génie des Procédés, Université des Sciences et de la Technologie d’Oran - Mohamed BOUDIAF, BP 1505 El M’naouer – Oran, Algérie; Architecture department, Laboratoire de Chimie des Matériaux Inorganiques et Applications, Université des Sciences et de la Technologie d’Oran - Mohamed BOUDIAF, BP 1505 El M’naouer – Oran, Algérie; Civil engineering department, Université des Sciences et de la Technologie d’Oran - Mohamed BOUDIAF, BP 1505 El M’naouer – Oran, Algérie

**Keywords:** Covid-19, Temperature, Humidity, Incubation period

## Abstract

Several studies have confirmed the impact of weather conditions on the evolution of the Covid-19 pandemic. We wanted to verify this phenomenon in the city of Oran in Algeria, which experienced its first case of Covid19 on March 19, 2020.

The data studied are the new Covid19 cases, the average, minimum and maximum temperatures, as well as the relative humidity rate.

A first analysis of the data with a Spearman rank correlation test did not yield significant results. Taking into account the average incubation period to adjust the data made it possible, during a second analysis, to show that the minimum temperature is significantly correlated with the new cases of Covid19 in Oran. This study can help establish prevention policies against Covid19, especially during fall in temperatures in autumn and winter.

## Introduction

The transmission of viruses can be impacted by several factors, including climatic conditions such as temperature and humidity as well as population density [1]. However, there is not yet enough time to fully understand the influence of meteorological parameters on the spread of the Covid19 virus. But already, several research works have been carried out on this field [2, 3, 4, 5] since the World Health Organization (WHO) declared that the Covid19 pandemic is a Public Health Emergency of International Concern (PHEIC), on January 30, 2020.

Algeria, like other countries in the world, has not escaped this Covid pandemic19 [6]. The first case reported on February 25, 2020, was imported from Italy [7]. In order to limit exposure to the virus, partial containment measures were imposed on certain cities that reported the highest number of contaminated cases. More and more people are invited to isolate themselves or to quarantine themselves, they only leave their homes to buy essential items such as food and medicines [8].

### Covid-19 at Oran

The city of Oran, the second largest city in Algeria, registered on March 19, 2020 its first case of Covid19. Figure 1 shows the progression of proven cases from Covid19 to Oran since that date.

**Fig. 1.**
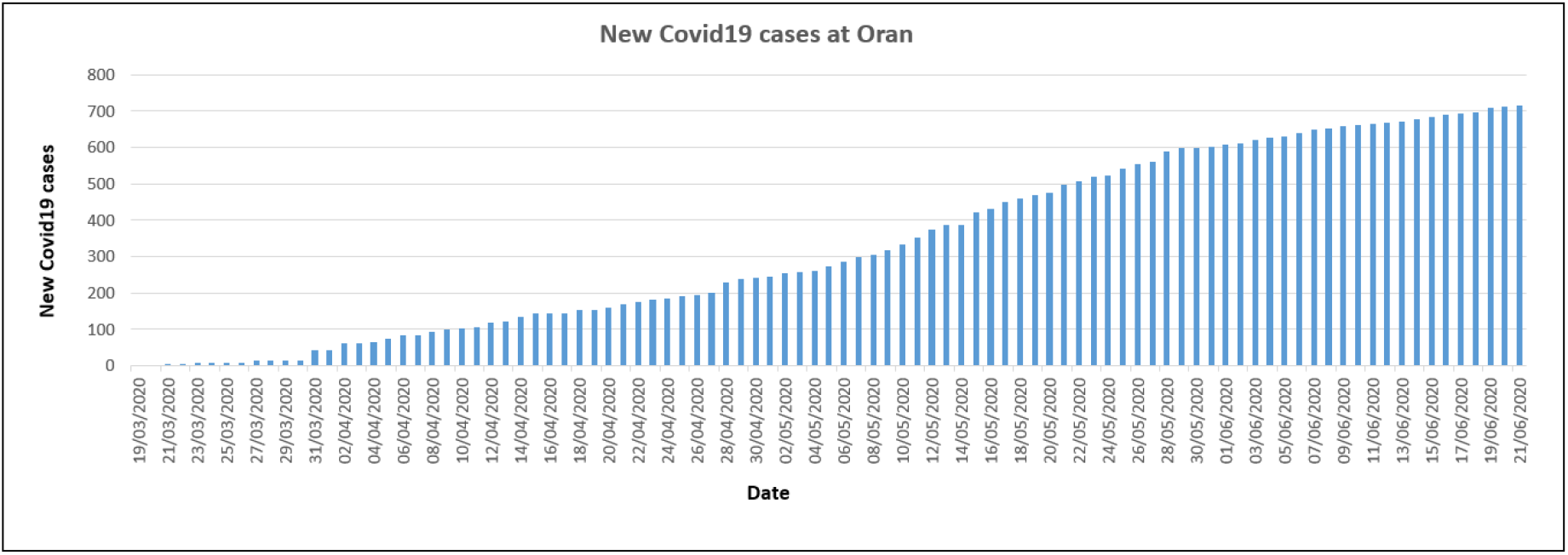
Number of confirmed cases infected with Covid-19 in Oran since March 19, 2020. **Source** : [9] ; compiled by authors.

Several models can help monitor an outbreak and predict its evolution. The most classic being the SIR model [10] which is based on the work of the theory of Kermack et al. [11]. We carried out a simulation of the SIR model for the period from 03/19/2020, date when the first case was registered in Oran until 04/04/2020, date of application of the strictest partial containment in Oran.

The result of the simulation gives a basic reproduction number R0 = 2.125638 for the above-mentioned period. Thus, at the start of the outbreak, a person infected with Covid19 could infect more than 2 people. This situation could have caused an exponential spread of the virus after 02 weeks as shown by the simulation of the SIR model presented in Figure 2. We can also see that this reproduction of the virus was probably slowed down by the sanitary and containment measures taken by Public powers. Indeed, containment is a barrier measure used to break the chains of virus transmission during an outbreak [12].

**Fig. 2.**
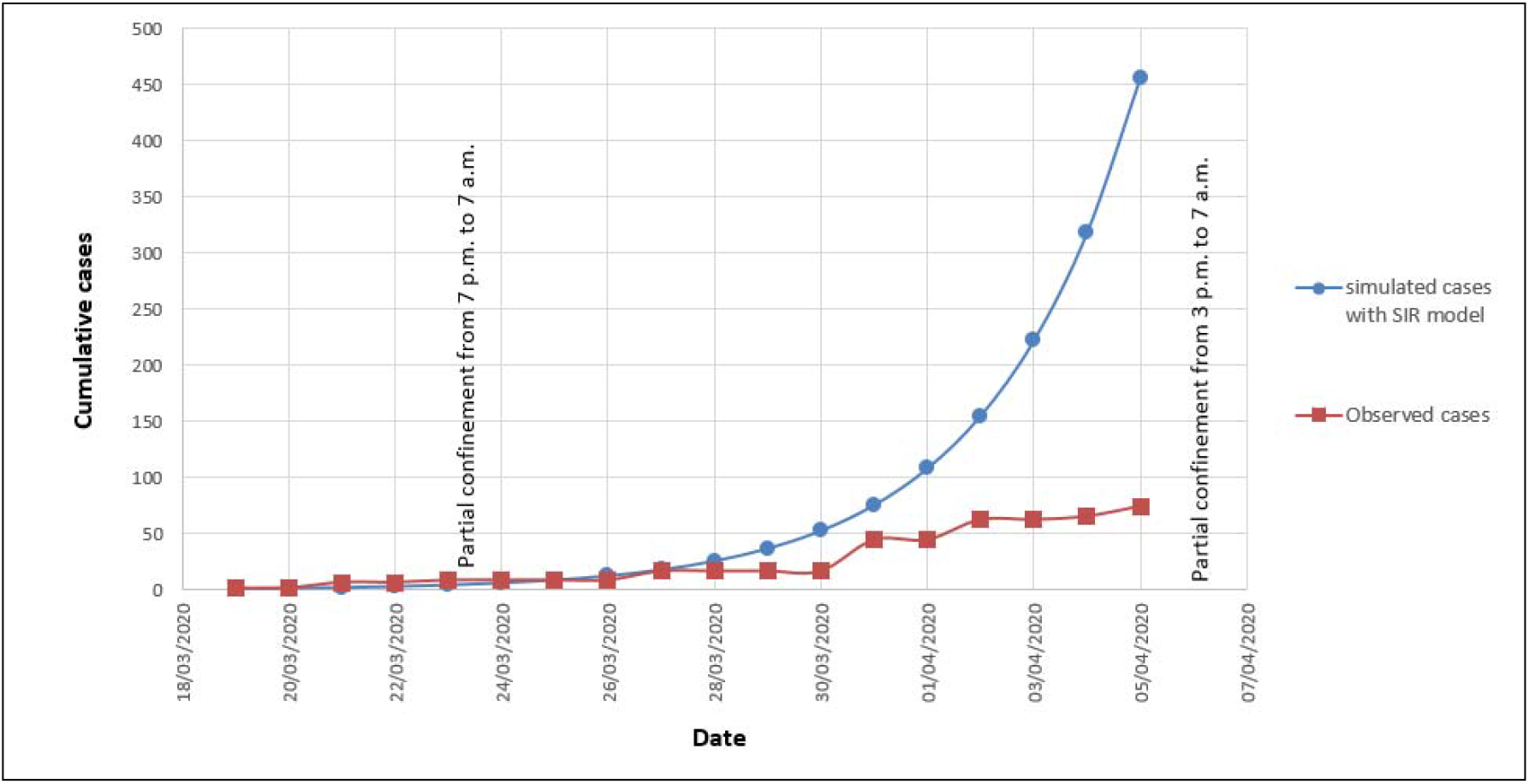
Comparison between simulated cases with SIR model and observed cases recorded by the MSPRH during the period from 19/03/2020 to 05/04/2020. **Source** : Authors.

### Impact of weather parameters on the propagation of Covid19 at Oran

The Oran region is characterized by a semi-arid climate [13] with annual rainfall less than 330 mm, occurring mainly between the months of October and May; average monthly temperatures vary between 5 ° and 17 ° C in winter and 16 ° and 31 ° C in summer [14]. The city of Oran is located in the northwest of Algeria, bordered by the Mediterranean Sea to the north and the Sebkha to the southwest as shown in Figure 3.

**Fig. 3.**
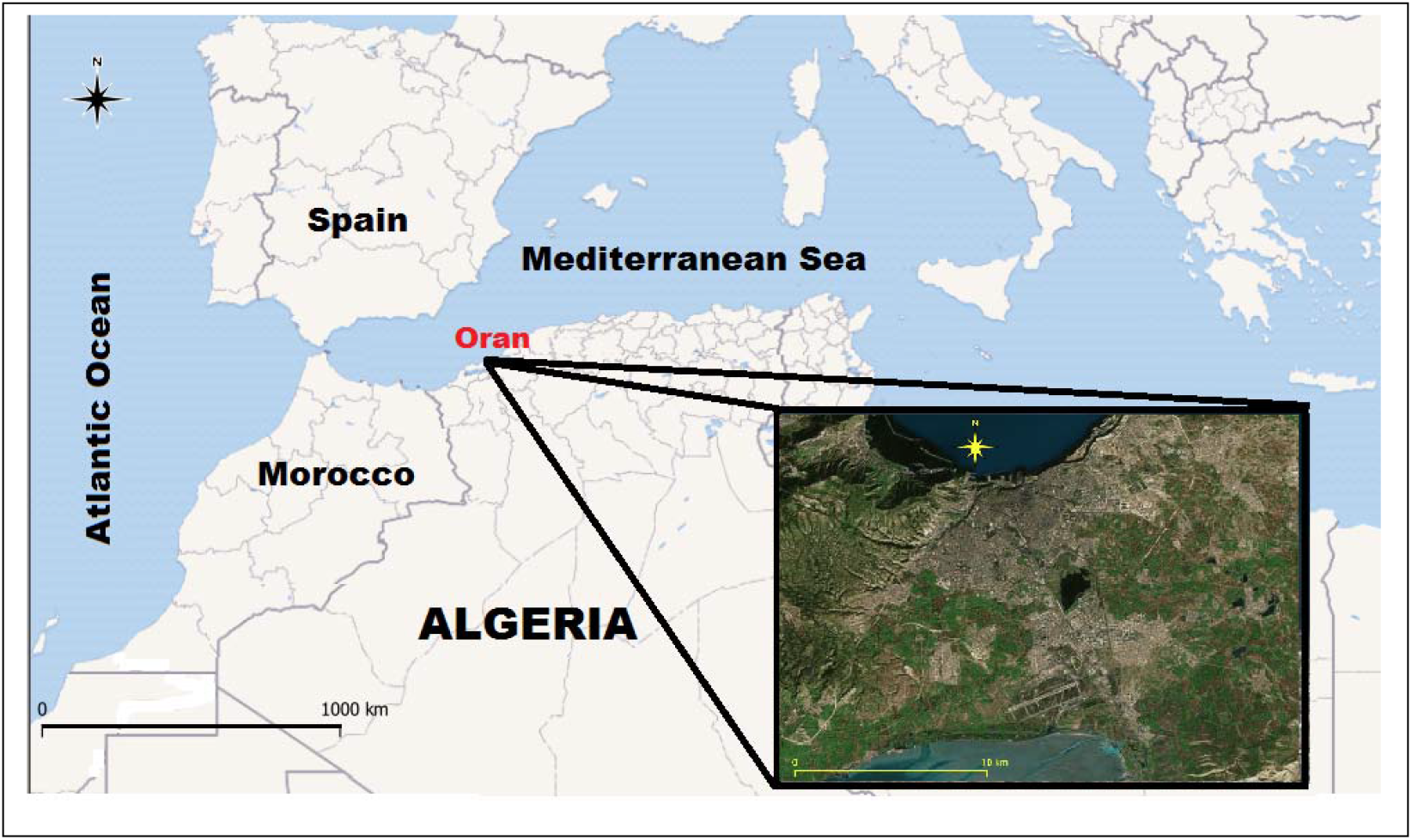
Geographical position of the city of Oran. **Source** : Authors.

Bukhari et al. [15] have shown that low temperature and humidity levels are key variables in determining the transmission of Covid19. Therefore, it is interesting to study this phenomenon for the region of Oran. For this study, we used the meteorological data recorded, during the months of April, May and June, at the meteorological station of Es Senia concerning minimum temperature (° C), average temperature (° C), maximum temperature (° C), and the relative humidity (%). Data on new Covid19 cases in Oran are from the MSPRH [9]. The evolution of temperature and humidity during the studied period is shown in Figure 4.

**Fig. 4.**
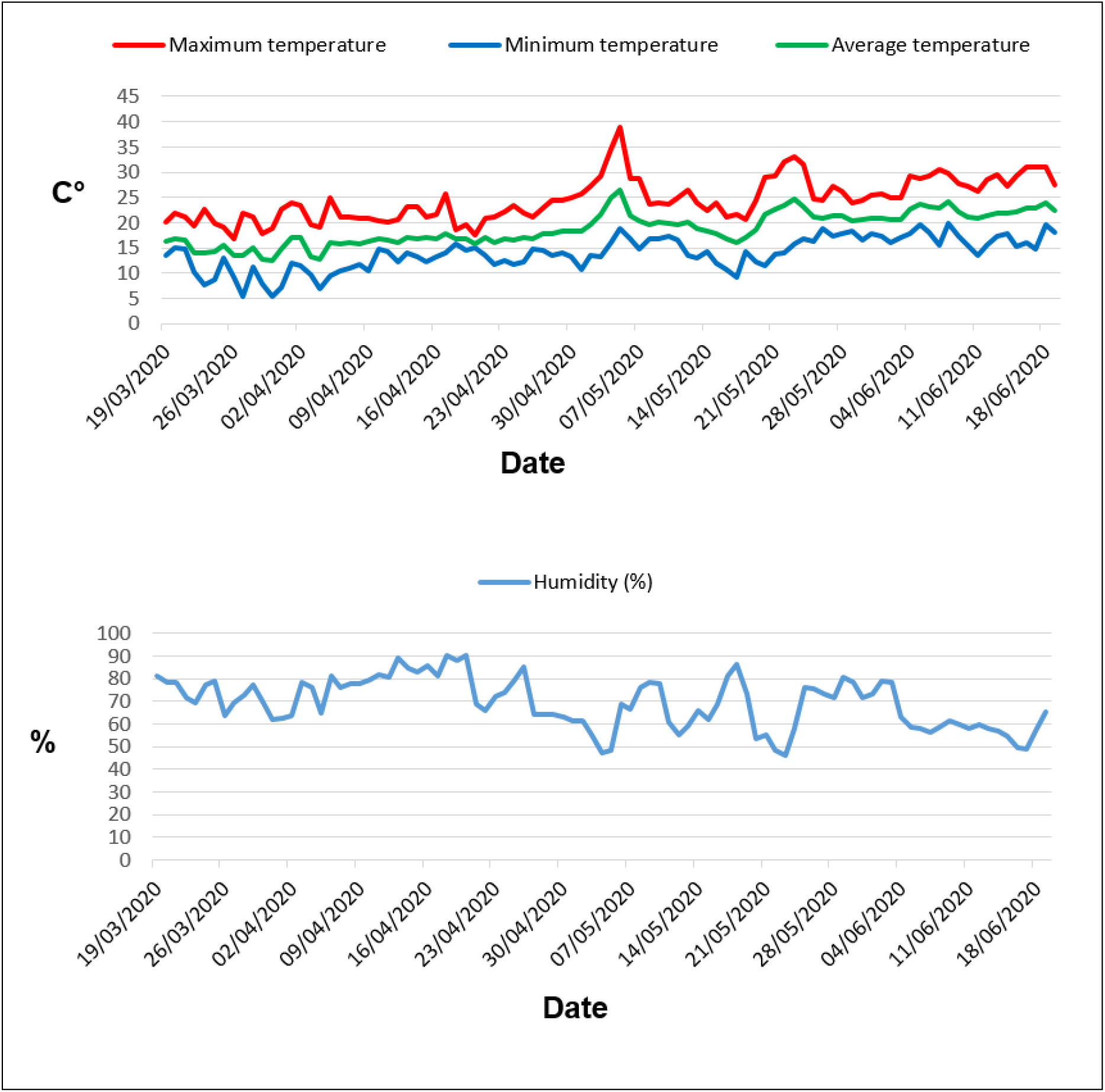
Evolution of temperature and humidity during the studied period, at the city of Oran. **Source** : Authors

A Spearman rank correlation test was used to analyze this data. The table 1 shows the obtained coefficients.

**Table 1.**
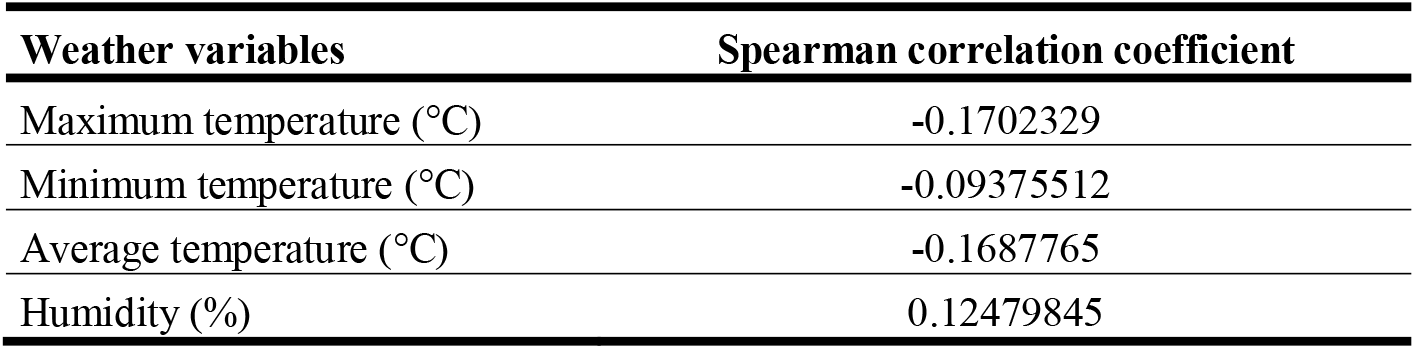
Spearman correlation coefficients between Covid-19 and weather variables.

| Weather variables | Spearman correlation coefficient |
| --- | --- |
| Maximum temperature (°C) | -0.1702329 |
| Minimum temperature (°C) | -0.09375512 |
| Average temperature (°C) | -0.1687765 |
| Humidity (%) | 0.12479845 |

The result does not show a significant correlation between the weather variables and the Covid19 in Oran. However, taking into account the incubation period of Covid19 has produced a significant result. Indeed, according to Wang et al. [16], the average incubation time of Covid19 is approximately 6.4 days. A 6-day shift in weather data from Covid19 data found a significant correlation between new Covid19 cases and minimum temperatures as shown in Table 2.

**Table 2.** Spearman correlation coefficients between Covid-19 and weather variables, taking into account the incubation period.

| Weather variables | Spearman correlation coefficient |
| --- | --- |
| Maximum temperature (°C) | 0.01290379 |
| Minimum temperature (°C) | -0.3123818 |
| Average temperature (°C) | -0.1086342 |
| Humidity (%) | -0.1411739 |

During the studied period, the minimum temperatures varied between 5.3 ° C and 19.9 ° C. The result confirms that temperature is an environmental driver of the Covid-19 outbreak as observed in China by Shi et al. [17].

## Conclusion

Even if the Covid19 pandemic in Oran seems relatively under control, the significant correlation between the minimum temperatures and the spread of the virus should alert health authorities to the risk of the epidemic worsening when the temperatures drop.

Given that meteorological parameters have a significant and consistent distribution of the seasonal behavior of respiratory viruses [18], it is desirable that the fight against Covid19 be intensified during the summer in order to anticipate a possible upsurge contaminations that could appear in autumn and winter.

## Declarations

### Availability of data and materials

All data generated or analyzed during this study can be obtained from the corresponding author.

## Data Availability

Data is on the websites below

http://covid19.sante.gov.dz/carte

https://worldweather.wmo.int

## Competing interests

The authors declare they have no competing interests.

## Funding

The authors confirm that no funding was received to carry out this study.

## Authors’ contributions

All the authors have contributed to the structure, content, and writing of the paper. All authors read and approved the final manuscript.

## Notes

### Competing Interest Statement

The authors have declared no competing interest.

### Funding Statement

No external funding was received

### Author Declarations

No approval need

